# Urinary 27-nor-5β-cholestane-3α,7α,12α,24R,25S-pentol glucuronide as a biomarker of non-small cell lung cancer in never-smokers: two independent case–control studies

**DOI:** 10.64898/2026.07.27.26359037

**Authors:** Seyon Chung, Huaitian Liu, Mohammed Khan, Tanvi S. Patel, Burchelle Blackman, Rolf E. Swenson, Sharon R. Pine, Frank J. Gonzalez, Curtis C. Harris, Daxesh P. Patel

## Abstract

**Background:** Lung cancer in never-smokers is excluded from low-dose computed tomography screening because eligibility is defined by smoking history, and no non-invasive marker is available to select never-smokers for imaging. We evaluated urinary 27-nor-5β-cholestane-3α,7α,12α,24*R*,25*S*-pentol glucuronide (CPG), a structurally authenticated bile alcohol glucuronide, as a marker of non-small cell lung cancer (NSCLC) in never-smokers.

**Methods:** CPG was quantified by ultra-performance liquid chromatography–tandem mass spectrometry against a synthetic standard in urine from two independent case–control studies (661 cases, 690 controls). The primary analyses compared never-smoker cases directly with never-smoker controls (NCI-Maryland, 114 cases and 203 controls; Colorado, 80 cases and 108 controls) by logistic regression adjusted for age, sex and race, for all stages and by stage, pooled across studies. Survival among never-smoker cases was analyzed by Kaplan–Meier and Cox regression. Analyses of all participants and of tumor transcriptomes were secondary.

**Results:** CPG was higher in never-smoker cases than in never-smoker controls in both studies (both P < 0.001). Adjusted odds ratios (OR) per standard deviation were near-identical across studies (1.83, 95% CI 1.32–2.54, and 1.81, 1.24–2.65; heterogeneity P = 0.96; pooled 1.82, 1.43–2.31). Elevation was present in stage I–II disease (pooled OR 1.38, 1.04–1.82) and greater in stage III–IV disease (2.27, 1.66–3.12). Single-marker discrimination was modest (area under the curve 0.63 and 0.70). High CPG was associated with shorter overall survival among never-smoker cases in NCI-Maryland independently of stage (hazard ratio 2.56, 1.58–4.14), and in the same direction in Colorado (1.63, 0.72–3.69). Among all participants, CPG was also higher in cases (pooled OR 1.61, 1.42–1.82), without effect modification by smoking, and remained associated after mutual adjustment for creatine riboside and *N*-acetylneuraminic acid. Tumors from patients with high urinary CPG showed enrichment of xenobiotic-metabolism, aryl hydrocarbon receptor/cytochrome P450 and Nrf2 programs.

**Conclusions:** Urinary CPG is reproducibly elevated in never-smokers with NSCLC compared with never-smoker controls, including in early-stage disease, in two independent studies. These phase 2 data support evaluation of CPG within multimarker panels in pre-diagnostic urine from never-smokers, a population with no current screening pathway.

## Background

Lung cancer is the most commonly diagnosed cancer and the leading cause of cancer death worldwide [1]. Although most cases are attributable to smoking, lung cancer in people who have never smoked would rank among the leading causes of cancer death if considered on its own, and its share of cases is rising, preferentially among women and East Asian populations [2, 3]. It is increasingly recognized as a distinct entity, with characteristic driver alterations and environmental risk factors, including air pollution, that differ from smoking-associated disease [3–5].

Low-dose computed tomography (LDCT) screening reduces lung cancer mortality in people with substantial smoking histories [6, 7]. Because eligibility is defined by pack-year exposure, never-smokers are excluded by design and have no approved screening pathway [8, 9], even though LDCT detects early-stage disease in selected never-smokers at rates comparable with those in smokers [10]. The limiting factor is therefore not the imaging test but the absence of a way to decide whom to image, and risk models built on smoking exposure cannot fill that gap. Non-invasive, biologically interpretable markers that could enrich risk in never-smokers are needed.

We previously reported urinary creatine riboside (CR) and *N*-acetylneuraminic acid (NANA) as markers of lung cancer in never-smokers and smokers in the same study populations analyzed here [11]. These metabolites report tumor-intrinsic metabolism. Bile-acid metabolism offers a complementary, host-side axis: hepatic bile-acid conjugation and glucuronide clearance are coupled to aryl hydrocarbon receptor (AhR) and cytochrome P450 (CYP) xenobiotic metabolism, the system through which environmental exposures that promote lung adenocarcinoma in never-smokers are processed [5, 12, 13].

27-nor-5β-cholestane-3α,7α,12α,24*R*,25*S*-pentol glucuronide (CPG), the glucuronide of a C_26_ bile alcohol, was first detected as an unidentified ion among the features that discriminated cancer status in the untargeted urinary screen that identified CR [14]. It has since been structurally solved and synthesized, together with a deuterated analog, enabling quantification against an authentic standard [15]. The aglycone excreted by patients with lung cancer carries 24*R*,25*S* stereochemistry, differing from the 24*S*,25*R* bile alcohol that predominates in healthy urine; because only the 24*R* tetrol is normally converted onward to cholic acid, its accumulation and glucuronide excretion are consistent with a disease-associated block in the 25-hydroxylation pathway of cholic acid biosynthesis [15–17].

Here we quantify urinary CPG in never-smokers from two independent case–control studies. Comparing never-smoker cases directly with never-smoker controls excludes tobacco exposure by design; within this comparison we examine the association by stage and its relationship with survival. As secondary analyses, we assess the association in all participants, including whether smoking modifies it, and describe the tumor expression programs associated with urinary CPG.

## Methods

### Study populations

Study populations, specimen collection and ethical approvals have been described previously [11]. The exploratory NCI-Maryland study enrolled 375 patients with NSCLC and 471 population controls from the greater Baltimore, Maryland, area between August 1997 and June 2019. Population controls were identified from Department of Motor Vehicles lists and frequency-matched to cases on age, sex and self-reported race, with preference for controls aged 60 years or older and a balanced sex distribution; controls who developed cancer within 5 years of recruitment were excluded, and cases had no other cancer diagnosis. Urine was collected in sterile 50 mL containers, divided into 10 mL aliquots at the University of Maryland and stored at −80 °C. The validation study enrolled 286 patients with NSCLC and 219 population controls at the University of Colorado; urine was collected in 60 mL specimen cups and stored at −80 °C. Samples were thawed on wet ice before analysis. Colorado participant numbers, including the smoking-status distribution of controls, differ slightly from those in the earlier report of this cohort (285 cases and 220 controls, of whom 79 and 111 were never-smokers) [11].

Never-smokers were defined as individuals who had smoked fewer than 100 cigarettes in their lifetime, and ever-smokers as former or current smokers. Never-smokers comprised 114 cases and 203 controls in NCI-Maryland and 80 cases and 108 controls in Colorado and formed the primary analysis population; analyses of all participants were secondary. Tumors were staged by a pathologist according to the seventh edition of the American Joint Committee on Cancer staging manual, and stage was grouped as early (I–II) or late (III–IV). Survival time was calculated from diagnosis to death or last follow-up, and deaths were ascertained through the National Death Index [11]; Colorado vital status was updated by National Death Index linkage. No formal sample-size calculation was performed.

### Urinary CPG quantification

Urine samples were prepared by protein precipitation, and CPG was quantified by ultra-performance liquid chromatography–tandem mass spectrometry (UPLC–MS/MS) on an ACQUITY UPLC BEH C18 column coupled to a Xevo TQ-S micro triple-quadrupole mass spectrometer (Waters, Milford, MA, USA) in positive electrospray multiple-reaction monitoring (*m/z* 615.40 → 385.33). CPG concentrations (nM) were quantified against a calibration curve prepared from the synthesized authentic 24*R*,25*S* CPG standard (4.5–10000 nM; lower limit of quantification, 4.5 nM) [15], with cortisol sulfate-d4 as internal standard, and were normalized to urinary creatinine measured by the Jaffe method [18]. Identity in urine was confirmed by matched retention time against the authentic standard (Additional file 1: Figure S1). Samples from the two studies were analyzed in separate batches, so all analyses used study-specific scales. Full assay conditions are given in Additional file 1.

### Tumor transcriptomics

RNA sequencing was performed on 83 samples (47 tumors and 36 matched adjacent non-tumor tissues) from 46 patients, as described in Additional file 1. Gene-level counts were converted to log_2_ counts per million, retaining genes with more than 1 count per million in at least 25% of samples. Tumors with matched urinary CPG were dichotomized at the median, and gene-set enrichment analysis (GSEA) [19] was performed on the genome-wide signed ranking of differential expression between high- and low-CPG tumors, against a curated collection of xenobiotic-metabolism, AhR/CYP, Nrf2 oxidative-stress and inflammatory gene sets and the MSigDB Hallmark collection [20]. As a sensitivity analysis, GSEA was repeated with CPG modeled as a continuous variable, and an AhR/CYP xenobiotic module score (mean *z*-scored expression of member genes) was related to continuous urinary CPG.

### Statistical analysis

The primary analyses compared never-smoker cases with never-smoker controls. CPG was compared between groups by the two-sided Mann–Whitney *U* test, and comparisons of never-smoker controls with early-stage and late-stage never-smoker cases were Holm-adjusted. For regression analyses, log_10_ CPG was standardized within each study, so odds ratios (OR) are expressed per standard deviation (SD). Logistic regression for case status was adjusted for age, sex and race and was fitted for all stages and separately for early- and late-stage disease. Pooled models added a study term, and between-study heterogeneity was assessed by Cochran’s *Q* test. Discrimination was summarized by the area under the receiver operating characteristic curve (AUC).

For survival, cases were dichotomized at the study-specific median CPG of all cases, and overall survival among never-smoker cases was analyzed by Kaplan–Meier estimation with log-rank tests and by Cox proportional-hazards regression in cases with complete staging, unadjusted and adjusted for stage, age and sex. The proportional-hazards assumption for CPG was met (Schoenfeld residual test, P = 0.34 and P = 0.15). Median follow-up was estimated by the reverse Kaplan–Meier method.

Secondary analyses in all participants used the same methods with additional adjustment for smoking status (never vs ever). Effect modification by smoking was tested with a CPG × smoking interaction term. Across smoking strata (never-smoker and ever-smoker controls and cases), the Kruskal–Wallis test was followed by four prespecified pairwise Mann–Whitney comparisons with Holm adjustment. CPG and CR + NANA, each dichotomized at the study-specific median as described previously [11], were mutually adjusted in logistic models adjusted for age, sex, race and smoking status, and Cox models in all cases were additionally adjusted for smoking status. In a sensitivity analysis, case–control and survival analyses were repeated after excluding four Colorado participants (two cases and two controls) whose case–control classification was uncertain in the study records. Analyses used Python 3.12 (SciPy, statsmodels, lifelines) and R version 4.0.5. All tests were two-sided, and P < 0.05 was considered statistically significant. Reporting follows the REMARK and STROBE recommendations [21, 22].

### Use of artificial intelligence tools

During the preparation of this manuscript the authors used Claude Sonnet 4.6 (Anthropic, San Francisco, CA, USA) for grammar and language editing only. No data, analysis or interpretation was generated by the tool. The authors subsequently reviewed and edited the content and take full responsibility for the content of the publication.

## Results

### Participant characteristics

Characteristics of never-smoker participants are shown in Table 1, and those of all participants in Additional file 1: Table S1. Never-smoker cases were predominantly women in both studies (70.2% and 65.0%). Never-smoker cases and controls differed in race distribution in NCI-Maryland (3.5% vs 47.8% African American) and in sex distribution in Colorado (35.0% vs 75.0% male); both were adjusted for in regression analyses. Late-stage disease accounted for 54.2% and 41.2% of staged never-smoker cases.

**Table 1.** Characteristics of never-smoker participants in the NCI-Maryland and Colorado studies.

| Characteristic | NCI-Maryland controls (n = 203) | NCI-Maryland cases (n = 114) | Colorado controls (n = 108) | Colorado cases (n = 80) |
| --- | --- | --- | --- | --- |
| Age, years, mean (SD) | 66.5 (9.5) | 64.7 (13.6) | 68.0 (13.6) | 66.1 (11.7) |
| **Sex, n (%)** |  |  |  |  |
| Male | 86 (42.4) | 34 (29.8) | 81 (75.0) | 28 (35.0) |
| Female | 117 (57.6) | 80 (70.2) | 26 (24.1) | 52 (65.0) |
| Missing | 0 | 0 | 1 (0.9) | 0 |
| **Race, n (%)** |  |  |  |  |
| European American<sup>a</sup> | 106 (52.2) | 110 (96.5) | 101 (93.5) | 72 (90.0) |
| African American<sup>a</sup> | 97 (47.8) | 4 (3.5) | 2 (1.9) | 1 (1.2) |
| Other or missing | 0 | 0 | 5 (4.6) | 7 (8.8) |
| **Stage, n (%)** |  |  |  |  |
| I | NA | 36 (31.6) | NA | 31 (38.8) |
| II | NA | 13 (11.4) | NA | 16 (20.0) |
| III | NA | 27 (23.7) | NA | 9 (11.2) |
| IV | NA | 31 (27.2) | NA | 24 (30.0) |
| Missing | NA | 7 (6.1) | NA | 0 |
| **Histology, n (%)** |  |  |  |  |
| Adenocarcinoma | NA | 104 (91.2) | NA | 74 (93.7) |
| Squamous cell carcinoma | NA | 10 (8.8) | NA | 5 (6.3) |
| Deaths, n (%) | NA | 81 (71.1) | NA | 27 (33.8) |
| Median follow-up, months<sup>b</sup> | NA | 116 | NA | 62 |
| Urinary CPG, nM, median (IQR) | 182 (93–359) | 292 (156–462) | 409 (300–675) | 652 (497–1031) |

Percentages are of all participants in each column. CPG, 27-nor-5β-cholestane-3 $\alpha$ ,7α ,12α ,24R,25S-pentol glucuronide; IQR, interquartile range; NA, not applicable; SD, standard deviation. <sup>a</sup>Self-reported. <sup>b</sup>Reverse Kaplan–Meier method.

### Urinary CPG is elevated in never-smoker cases, including early-stage disease

Among never-smokers, CPG was higher in cases than in controls in NCI-Maryland (median 292 vs 182 nM; P < 0.001) and Colorado (652 vs 409 nM; P < 0.001). Elevation was present in early-stage disease in both studies (Holm-adjusted P = 0.04 and P = 0.002) and was greatest in late-stage disease (both P < 0.001; Fig. 1). AUCs for never-smoker cases versus never-smoker controls were 0.63 in NCI-Maryland and 0.70 in Colorado, and 0.60 and 0.66 for early-stage disease.

**Fig. 1.**
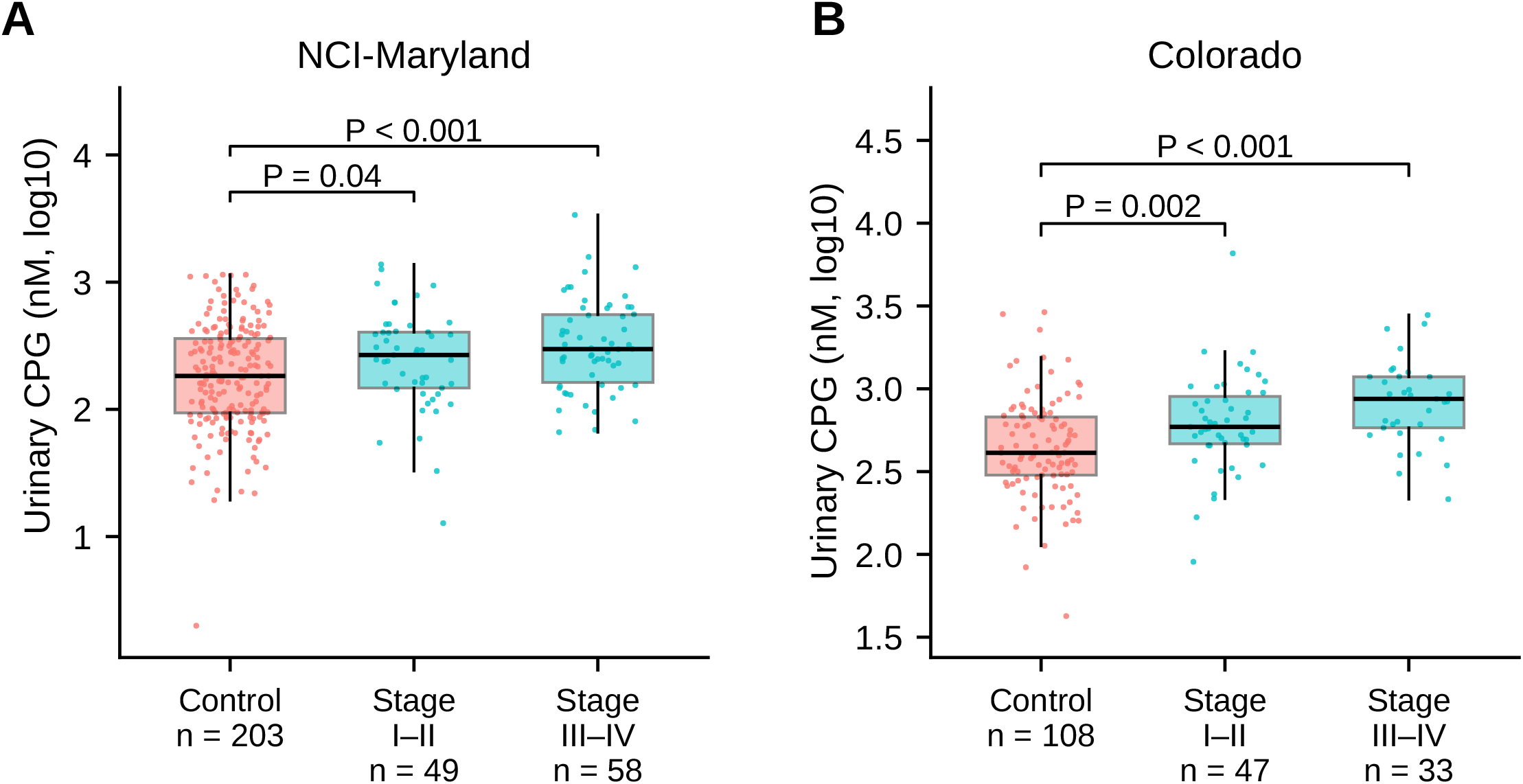
Urinary CPG in never-smokers by stage group in two independent studies. Urinary CPG (nM, log_10_ scale) in never-smoker controls and never-smoker cases with stage I–II and stage III–IV non-small cell lung cancer in the **A** NCI-Maryland and **B** Colorado studies. Boxes show the median and interquartile range, whiskers extend to 1.5 times the interquartile range, and individual participant values are overlaid. Group sizes are given beneath each box. Brackets show two-sided Mann–Whitney *U* tests versus controls, Holm-adjusted for two comparisons. Seven NCI-Maryland never-smoker cases with missing stage are not shown.

### The adjusted association is replicated across studies

After adjustment for age, sex and race, the association in never-smokers was almost identical in the two studies: OR per SD 1.83 (95% CI 1.32–2.54) in NCI-Maryland and 1.81 (1.24–2.65) in Colorado, with no evidence of heterogeneity (P = 0.96) and a pooled OR of 1.82 (1.43–2.31) (Table 2; Fig. 2). For early-stage disease, adjusted ORs were 1.46 (0.99–2.16) and 1.24 (0.82–1.88), without heterogeneity (P = 0.57), and the pooled estimate was statistically significant (OR 1.38, 95% CI 1.04–1.82; P = 0.02). Associations were stronger in late-stage disease (pooled OR 2.27, 95% CI 1.66–3.12).

**Table 2.** Associations of urinary CPG with case status and overall survival among never-smokers.

| Analysis | NCI-Maryland | Colorado | Pooled | Heterogeneity P value |
| --- | --- | --- | --- | --- |
| **Case status, adjusted OR per SD (95% CI)** |  |  |  |  |
| All stages<sup>a</sup> | 1.83 (1.32– 2.54) | 1.81 (1.24– 2.65) | 1.82 (1.43– 2.31) | 0.96 |
| Stage I–II<sup>a</sup> | 1.46 (0.99– 2.16) | 1.24 (0.82– 1.88) | 1.38 (1.04– 1.82) | 0.57 |
| Stage III–IV<sup>a</sup> | 2.15 (1.41– 3.28) | 2.49 (1.49– 4.18) | 2.27 (1.66– 3.12) | 0.67 |
| **Overall survival, HR high vs low CPG (95% CI)** |  |  |  |  |
| Unadjusted<sup>b</sup> | 2.16 (1.35– 3.46) | 2.15 (1.00– 4.64) | NA | NA |
| Adjusted for stage, age and sex<sup>b</sup> | 2.56 (1.58– 4.14) | 1.63 (0.72– 3.69) | NA | NA |

**Fig. 2.**
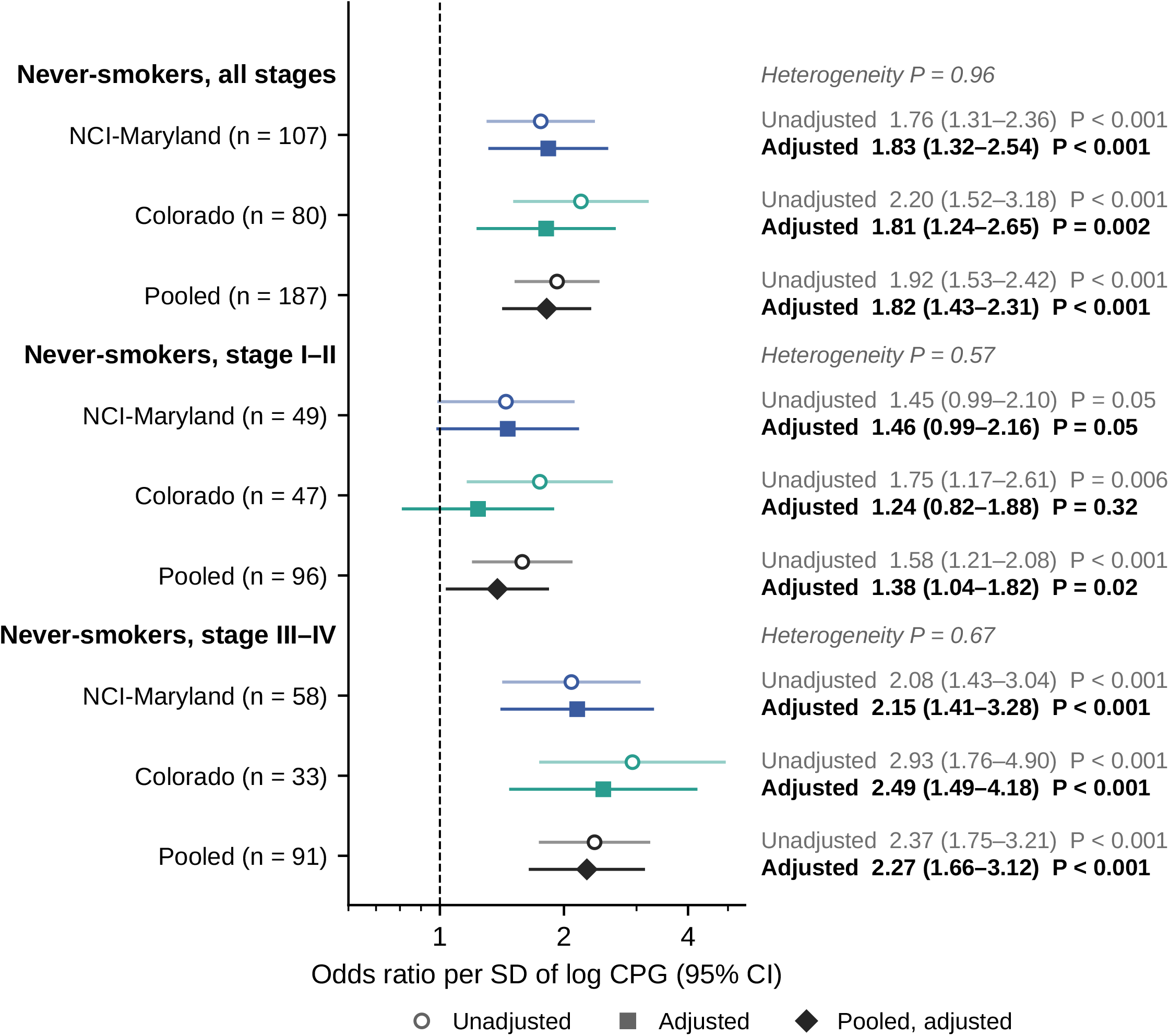
Odds ratios for case status among never-smokers by stage group. Odds ratios per standard deviation of within-study standardized log_10_ CPG (nM) for never-smoker cases versus never-smoker controls, for all stages, stage I–II and stage III–IV disease; n is the number of cases with known stage. Open circles, unadjusted estimates; squares, estimates adjusted for age, sex and race; diamonds, pooled adjusted estimates including a study term. Horizontal lines are 95% confidence intervals, and the dashed line marks an odds ratio of 1 on a logarithmic scale. Heterogeneity P values are from Cochran’s *Q* test. All tests were two-sided.

In Colorado, adjustment attenuated the early-stage estimate from 1.75 to 1.24. This reflected sex imbalance: 75% of never-smoker controls but 35% of never-smoker cases were men, and CPG was higher in women than in men among controls (P = 0.01).

### CPG and overall survival in never-smoker cases

Among never-smoker cases, high CPG was associated with shorter overall survival in NCI-Maryland (log-rank P = 0.002; Fig. 3A). In 107 never-smoker cases with complete staging (76 deaths), the association strengthened after adjustment for stage, age and sex (unadjusted hazard ratio [HR] 2.16, 95% CI 1.35–3.46; adjusted HR 2.56, 95% CI 1.58–4.14), and high- and low-CPG cases had similar proportions of late-stage disease (57% vs 52%). In Colorado (80 never-smoker cases, 27 deaths), the unadjusted association was in the same direction and of similar magnitude (log-rank P = 0.045; HR 2.15, 95% CI 1.00–4.64; Fig. 3B) but was attenuated after adjustment (HR 1.63, 95% CI 0.72–3.69), with high-CPG cases more often having late-stage disease (58% vs 30%; Fig. 3C; Table 2).

**Fig. 3.**
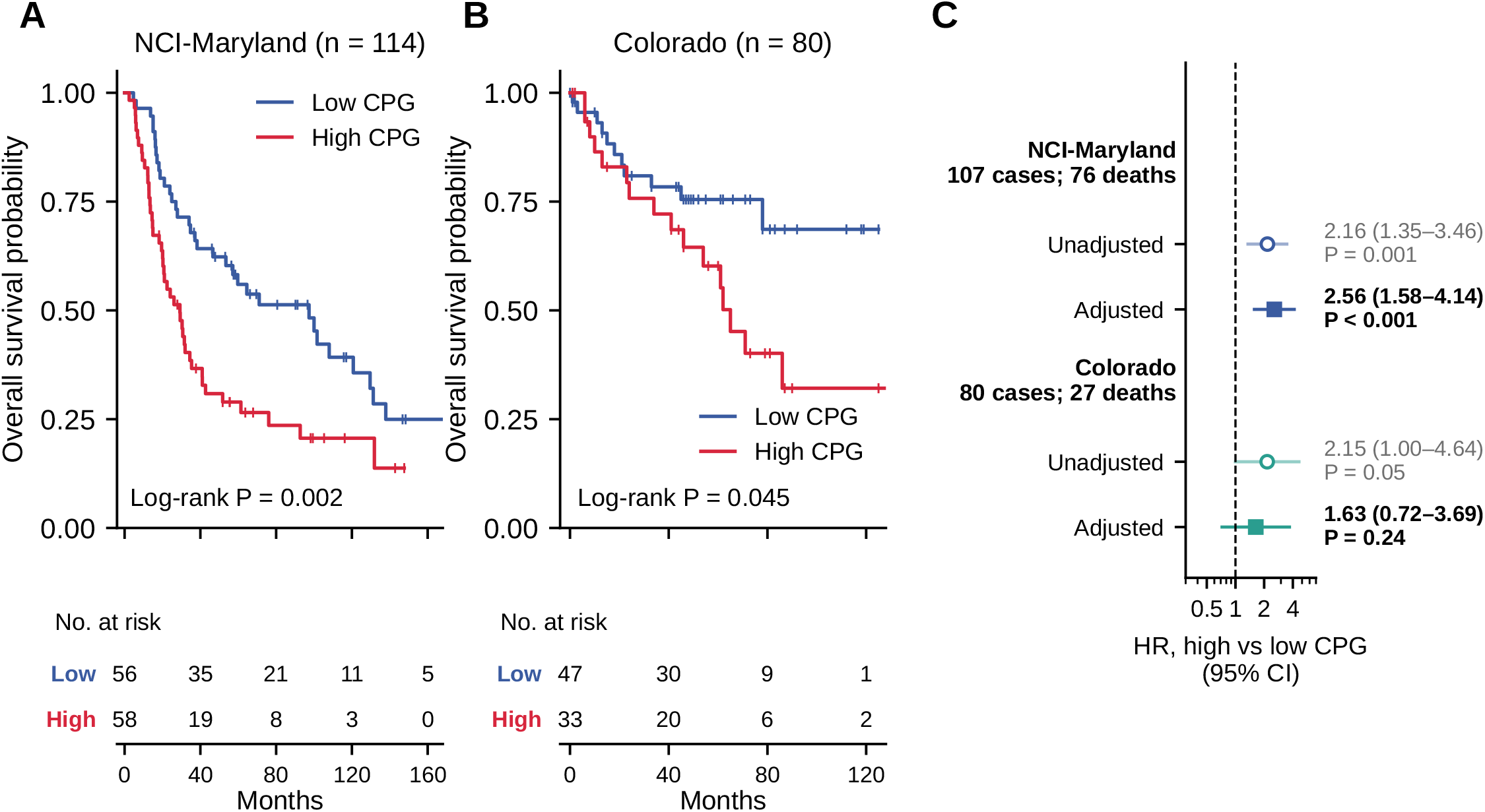
Overall survival among never-smoker cases by urinary CPG. Kaplan–Meier estimates of overall survival among never-smoker cases dichotomized at the study-specific median CPG of all cases in the **A** NCI-Maryland and **B** Colorado studies, with log-rank P values; tick marks indicate censoring and numbers at risk are shown beneath each panel. **C** Hazard ratios for high versus low CPG among never-smoker cases with complete staging; open circles, unadjusted; squares, adjusted for stage, age and sex. Horizontal lines are 95% confidence intervals, and the dashed line marks a hazard ratio of 1 on a logarithmic scale. Colorado vital status reflects National Death Index linkage. All tests were two-sided.

### Secondary analyses in all participants

Among all participants, CPG was higher in cases than in controls in NCI-Maryland (median 284 vs 213 nM; P < 0.001) and Colorado (825 vs 473 nM; P < 0.001; Additional file 1: Figure S2). After adjustment for age, sex, race and smoking status, the OR per SD was 1.48 (95% CI 1.27–1.72) in NCI-Maryland and 1.91 (1.52–2.39) in Colorado, with a pooled OR of 1.61 (1.42–1.82) (heterogeneity P = 0.07; Additional file 1: Table S2). There was no evidence that the association differed by smoking status (CPG × smoking interaction P = 0.21 and P = 0.94), and ever-smoker cases also exceeded ever-smoker controls (Holm-adjusted P = 0.04 and P < 0.001). CPG remained associated with case status after mutual adjustment for CR + NANA (adjusted OR for high vs low CPG 1.58, 95% CI 1.15–2.16, in NCI-Maryland and 3.92, 2.47–6.29, in Colorado), indicating that it is not a surrogate for the previously reported markers. Among all cases, HRs for high versus low CPG adjusted for stage, age, sex and smoking status were 1.53 (95% CI 1.21–1.94) in NCI-Maryland and 1.42 (0.96–2.11) in Colorado. AUCs for all cases versus all controls were 0.59 and 0.71.

### Sensitivity analysis

Excluding the four Colorado participants with uncertain case–control classification did not materially change the results (Additional file 1: Table S3): the Colorado adjusted OR per SD among never-smokers was 1.80 (95% CI 1.23–2.63), the pooled early-stage never-smoker OR was 1.37 (1.04–1.81), and the unadjusted HR among Colorado never-smoker cases was 2.29 (1.06–4.94).

### Tumor expression programs associated with urinary CPG

Tumors from patients with high urinary CPG showed positive enrichment of xenobiotic phase I/II, xenobiotic-metabolism, tobacco-smoke-metabolism, Nrf2 oxidative-stress and AhR/CYP core gene sets, including the CYP1A1/CYP1B1 AhR axis (Additional file 1: Figure S3). This xenobiotic axis was reproduced when CPG was modeled as a continuous variable, whereas Hallmark and inflammatory gene-set associations were not robust to this analytic choice and are not interpreted (Additional file 1: Figure S4). An AhR/CYP xenobiotic module score showed a weak positive association with continuous urinary CPG (*R*^2^ = 0.05, P = 0.048; Spearman ρ = 0.21, P = 0.07; Additional file 1: Figure S5). Never-smokers contributed only a small minority of sequenced tumors, so these data characterize the xenobiotic axis across patients as a whole.

## Discussion

In two independent case–control studies, urinary CPG was elevated in never-smokers with NSCLC compared directly with never-smoker controls, with near-identical adjusted effect sizes in studies that differ in geography, ancestry and sex distribution. Because this comparison excludes tobacco exposure by design, the elevation cannot be attributed to smoking; consistent with this, the association was also present among all participants, with no evidence of effect modification by smoking. The elevation was present in early-stage disease in pooled analysis, with both studies pointing in the same direction, and it persisted after mutual adjustment for CR and NANA, indicating that CPG captures information not carried by those markers.

Three properties distinguish CPG from empirically derived candidates. First, it is chemically authenticated: the compound was structurally solved and synthesized, and quantification uses an authentic standard rather than an untargeted feature [15]. Second, it carries a disease-associated chemical signature: the 24*R*,25*S* configuration of the excreted sterol implies altered hepatic bile-acid handling in patients rather than a tumor-derived product [15, 16]. Third, the tumor expression data link high urinary CPG to coordinated AhR/CYP and Nrf2 xenobiotic programs, the system that processes environmental carcinogens implicated in never-smoker lung cancer [5, 12]. These observations are correlative and generate a testable hypothesis, namely that CPG reports a host–environment interface engaged in lung carcinogenesis, rather than establishing a mechanism.

The stage gradient indicates that CPG partly reflects disease extent, which is expected for a marker measured at diagnosis. The survival data are consistent with this but are not explained by it: in NCI-Maryland the hazard ratio among never-smokers increased after stage adjustment, and high- and low-CPG cases had similar stage distributions. Among all cases, adjusted hazard ratios were similar in the two studies (Additional file 1: Table S2). In Colorado never-smokers the unadjusted association was of similar magnitude but attenuated after adjustment, in a cohort with about one-third as many deaths; we therefore regard the prognostic association as supportive rather than validated.

Within the phase framework for biomarker development, this is a phase 2 study: assay development and case–control discrimination at clinical diagnosis in two independent populations [23, 24]. Case–control designs with prevalent, clinically detected cases overstate screening performance, so we report no sensitivity or specificity operating point and make no detection claim. Single-marker discrimination was modest, and any clinical use would be as one component of a panel with CR and NANA. The appropriate next step is evaluation in pre-diagnostic urine from prospective cohorts, under a prospective-specimen-collection, retrospective-blinded-evaluation design, and as a companion study within never-smoker LDCT programs, where imaging provides the reference standard [9, 10, 25].

This study has limitations. Specimens were collected at diagnosis and cannot establish how far in advance of diagnosis CPG rises. Never-smoker early-stage groups were small, so stage-specific estimates in each study were imprecise. Absolute CPG concentrations differed between studies, so a transferable threshold will require harmonized, calibrated measurement. Never-smoker cases and controls differed in race distribution in NCI-Maryland and in sex distribution in Colorado; although both were adjusted for, residual confounding by these or by diet, secondhand smoke, hepatic or renal function and other environmental exposures cannot be excluded. Never-smoking status was self-reported and was not verified by cotinine measurement. The transcriptomic cohort contained few never-smokers and was not adjusted for smoking status, and CPG has not yet been evaluated in serum or in East Asian populations, where the never-smoker burden is greatest.

## Conclusions

Urinary CPG, a chemically authenticated bile alcohol glucuronide, is reproducibly elevated in never-smokers with NSCLC compared with never-smoker controls, including in early-stage disease, in two independent studies, and is associated with tumor xenobiotic-metabolism programs. Prospective evaluation in pre-diagnostic urine from never-smokers, within multimarker panels, is warranted for a population that current screening criteria exclude.

## Data Availability

De-identified data supporting these findings are available within the article and its supplementary information, or from the corresponding author on reasonable request. RNA-sequencing gene-level counts and metadata are provided as supplementary data files. Raw data are generated at the National Cancer Institute, NIH.

## Abbreviations

AhR: Aryl hydrocarbon receptor
AUC: Area under the receiver operating characteristic curve
CI: Confidence interval
CPG: 27-nor-5β-cholestane-3α,7α,12α,24*R*,25*S*-pentol glucuronide
CR: Creatine riboside
CYP: Cytochrome P450
GSEA: Gene-set enrichment analysis
HR: Hazard ratio
IQR: Interquartile range
LDCT: Low-dose computed tomography
NANA: *N*-acetylneuraminic acid
NSCLC: Non-small cell lung cancer
OR: Odds ratio
SD: Standard deviation
UPLC–MS/MS: Ultra-performance liquid chromatography–tandem mass spectrometry

## Supplementary Information

**Additional file 1** (.docx). Supplementary Methods; Table S1, characteristics of all participants; Table S2, associations of urinary CPG with case status and survival among all participants; Table S3, sensitivity analysis excluding four Colorado participants; Figure S1, structural identification and chromatographic detection of urinary CPG; Figure S2, urinary CPG in all participants, overall and by smoking status; Figure S3, tumor gene-set enrichment by median-split urinary CPG; Figure S4, tumor gene-set enrichment with urinary CPG as a continuous variable; Figure S5, AhR/CYP xenobiotic module score versus urinary CPG.

## Declarations

### Ethics approval and consent to participate

The NCI-Maryland study (National Cancer Institute protocol OH98-C-N027; ClinicalTrials.gov NCT00339859) was approved by the Institutional Review Boards of the National Cancer Institute, National Institutes of Health, and of the seven participating recruitment institutions: the Baltimore Veterans Administration Medical Center, Bon Secours Hospital, MedStar Harbor Hospital, Sinai Hospital, Johns Hopkins Bayview Medical Center, The Johns Hopkins Hospital and the University of Maryland Medical Center. The Colorado study was approved by the University of Colorado Cancer Center PRMS Expedited Review Committee (PRMS 23-110) and the Colorado Multiple Institutional Review Board (COMIRB 23-0941). Written informed consent, or a waiver of consent, was obtained from all participants. The study was performed in accordance with the Declaration of Helsinki.

### Consent for publication

Not applicable.

### Availability of data and materials

The RNA-sequencing dataset supporting the conclusions of this article is available in the NCBI Gene Expression Omnibus repository, GSE344573, https://www.ncbi.nlm.nih.gov/geo/query/acc.cgi?acc=GSE344573. The untargeted urinary metabolomics dataset from the parent study is available in the MetaboLights repository, MTBLS28, https://www.ebi.ac.uk/metabolights/MTBLS28. The urinary CPG and clinical datasets used and analyzed during the current study are available from the corresponding author on reasonable request, subject to the data-sharing policies of the National Cancer Institute, National Institutes of Health.

### Authors’ contributions

S.C.: data curation, formal analysis, validation, software, writing (original draft). H.L.: formal analysis, validation, software, writing (original draft). M.K.: formal analysis, software, writing (review and editing). T.S.P.: formal analysis, software, writing (review and editing). B.B.: resources, writing (review and editing). R.E.S.: resources, writing (review and editing). S.R.P.: resources, writing (review and editing). F.J.G.: resources, writing (review and editing). C.C.H.: conceptualization, funding acquisition, investigation, project administration, resources, supervision, writing (review and editing). D.P.P.: conceptualization, data curation, investigation, methodology, project administration, supervision, writing (original draft). All authors read and approved the final manuscript.

## Acknowledgments

We thank the staff of the National Cancer Institute, the University of Maryland and the University of Colorado for clinical information, demographic data and biospecimens. The contributions of the National Institutes of Health authors were made as part of their official duties as NIH federal employees and are considered Works of the United States Government. The findings and conclusions presented in this paper are those of the authors and do not necessarily reflect the views of the NIH or the U.S. Department of Health and Human Services.

