## Additional File for "Urinary 27-nor-5β-cholestane-3α,7α,12α,24R,25S-pentol glucuronide as a biomarker of non-small cell lung cancer in never-smokers: two independent case–control studies"

**Additional file 1**

**Supplementary Methods**

**Sample preparation.** Urine samples were thawed on wet ice. For each sample, 60 µL of urine was combined with 60 µL of ice-cold acetonitrile:methanol:water (65:5:30, v/v/v) containing 2.5 nM of the internal standard cortisol sulfate-d4 to precipitate protein. Samples were mixed for 10 min and centrifuged at 14000 rpm for 15 min at 4 °C, and 100 µL of the clarified supernatant was transferred to an autosampler vial. The autosampler was maintained at 12 °C.

**Chromatography.** Separation used an ACQUITY UPLC BEH C18 column (1.7 µm, 2.1 × 50 mm; Waters, Milford, MA, USA) maintained at 40 °C. Mobile phase A was water with 0.1% formic acid and mobile phase B was acetonitrile with 0.1% formic acid, delivered at 0.400 mL/min. The injection volume was 5 µL and the total run time was 11.5 min. The linear gradient was 2% B held to 0.50 min, 20% B at 4.00 min, 95% B at 8.00 min and 99% B at 8.20 min, held to 9.00 min, then returned to 2% B at 9.10 min and re-equilibrated to 10.00 min. CPG eluted at 5.69 min.

**Mass spectrometry.** Analytes were detected on a Xevo TQ-S micro triple-quadrupole mass spectrometer (Waters) with positive electrospray ionization (capillary voltage 2.85 kV, source temperature 140 °C, desolvation temperature 400 °C, desolvation gas flow 1000 L/h, collision gas on). Data were acquired in multiple-reaction-monitoring mode with a mass span of 0.2 Da. CPG (C_32_H_54_O_11_) was monitored as the [M+H]^+^ precursor ion at *m/z* 615.40 (cone voltage 2 V) with the quantifier transition 615.40 → 385.33 (collision energy 20 V). The internal standard cortisol sulfate-d4 was monitored at *m/z* 447.07 (cone voltage 40 V) with the quantifier transition 447.07 → 367.16 (collision energy 12 V). Identity in urine was confirmed by matched retention time against the authentic standard (Figure S1).

**Quantification and normalization.** Peak areas were processed in MassLynx/TargetLynx (Waters) with multiple-reaction-monitoring spike removal enabled and smoothing disabled. CPG was quantified against a calibration curve prepared from the synthesized authentic 24*R*,25*S* CPG standard (4.5–10000 nM; lower limit of quantification, 4.5 nM), with cortisol sulfate-d4 as internal standard. CPG concentrations (nM) were normalized to urinary creatinine measured by the Jaffe method. Samples from the two studies were analyzed in separate batches; absolute concentrations therefore differ between studies, and all statistical analyses used study-specific scales.

**RNA sequencing and gene-set enrichment.** Gene-level counts were obtained for 83 samples (47 tumors and 36 matched adjacent non-tumor tissues) from 46 patients. Counts were converted to log_2_ counts per million with TMM normalization, retaining genes with more than 1 count per million in at least 25% of samples. For CPG-stratified analysis, tumors with matched urinary CPG were dichotomized at the median CPG, and differential expression was assessed with limma-voom. GSEA was performed on the genome-wide signed ranking of differential expression against MSigDB Hallmark gene sets and a curated collection of xenobiotic-metabolism, AhR/CYP, Nrf2 oxidative-stress and inflammatory gene sets. Normalized enrichment scores (NES) and Benjamini–Hochberg-adjusted P values are reported. Parallel analyses ranking on continuous urinary CPG used identical parameters. Curated AhR/CYP xenobiotic gene sets were scored as the mean *z*-scored normalized expression of member genes per sample and tested for association with continuous urinary CPG by linear regression and Spearman correlation.

**Additional statistical detail.** Log_10_ CPG (nM; 1 was added to accommodate zero values) was standardized to mean 0 and SD 1 within each study. Race was entered as a binary covariate (NCI-Maryland: European American vs African American; Colorado: European American vs other). Smoking status was entered as never vs ever. Between-study heterogeneity of adjusted log odds ratios was assessed by Cochran’s *Q* test with 1 degree of freedom. For survival analyses, cases were dichotomized at the median CPG of all cases in the respective study (284 nM in NCI-Maryland and 825 nM in Colorado). Cox models were fitted among cases with complete staging, with stage entered as I–II vs III–IV. The proportional-hazards assumption for CPG in never-smoker cases was assessed by the Schoenfeld residual test (P = 0.34 and P = 0.15). Median follow-up was estimated by the reverse Kaplan–Meier method.

**Supplementary Tables**

**Table S1** Characteristics of all participants in the NCI-Maryland and Colorado studies

| **Characteristic** | **NCI-Maryland controls**  **(n = 471)** | **NCI-Maryland cases**  **(n = 375)** | **Colorado controls**  **(n = 219)** | **Colorado cases**  **(n = 286)** |
| --- | --- | --- | --- | --- |
| Age, years, mean (SD) | 66.4 (9.1) | 65.7 (9.2) | 69.4 (12.2) | 69.1 (10.4) |
| **Sex, n (%)** |  |  |  |  |
| Male | 239 (50.7) | 181 (48.3) | 162 (74.0) | 118 (41.3) |
| Female | 232 (49.3) | 194 (51.7) | 56 (25.6) | 168 (58.7) |
| Missing | 0 | 0 | 1 (0.5) | 0 |
| **Race, n (%)** |  |  |  |  |
| European American^a^ | 257 (54.6) | 302 (80.5) | 197 (90.0) | 263 (92.0) |
| African American^a^ | 214 (45.4) | 73 (19.5) | 12 (5.5) | 6 (2.1) |
| Other or missing | 0 | 0 | 10 (4.6) | 17 (5.9) |
| **Smoking status, n (%)** |  |  |  |  |
| Never | 203 (43.1) | 114 (30.4) | 108 (49.3) | 80 (28.0) |
| Ever | 268 (56.9) | 261 (69.6) | 111 (50.7) | 206 (72.0) |
| Former | NA | 132 (35.2) | NA | 179 (62.6) |
| Current | NA | 129 (34.4) | NA | 27 (9.4) |
| **Stage, n (%)** |  |  |  |  |
| I | NA | 182 (48.5) | NA | 103 (36.0) |
| II | NA | 45 (12.0) | NA | 66 (23.1) |
| III | NA | 79 (21.1) | NA | 35 (12.2) |
| IV | NA | 58 (15.5) | NA | 80 (28.0) |
| Missing | NA | 11 (2.9) | NA | 2 (0.7) |
| **Histology, n (%)** |  |  |  |  |
| Adenocarcinoma | NA | 267 (71.2) | NA | 235 (82.5)^d^ |
| Squamous cell carcinoma | NA | 108 (28.8) | NA | 49 (17.2)^d^ |
| Deaths, n (%) | NA | 297 (79.2) | NA | 103 (36.0) |
| Median follow-up, months^b^ | NA | 219 | NA | 60 |
| Urinary CPG, nM, median (IQR)^c^ | 213 (100–386) | 284 (145–594) | 473 (311–751) | 825 (549–1239) |

Percentages are of all participants in each column. Former and current smoking were recorded for cases only in the analysis files. CPG, 27-nor-5β-cholestane-3α,7α,12α,24*R*,25*S*-pentol glucuronide; IQR, interquartile range; NA, not applicable; SD, standard deviation. ^a^Self-reported. ^b^Reverse Kaplan–Meier method.

**Table S2** Associations of urinary CPG with case status and overall survival among all participants

| **Analysis** | **NCI-Maryland** | **Colorado** | **Pooled** | **Heterogeneity**  **P value** |
| --- | --- | --- | --- | --- |
| **Case status, OR per SD (95% CI)** |  |  |  |  |
| Unadjusted | 1.39 (1.20–1.60) | 2.19 (1.77–2.72) | 1.62 (1.44–1.83) | NA |
| Adjusted^a^ | 1.48 (1.27–1.72) | 1.91 (1.52–2.39) | 1.61 (1.42–1.82) | 0.07 |
| CPG × smoking interaction, P^b^ | 0.21 | 0.94 | NA | NA |
| **Case status, OR high vs low CPG (95% CI)** |  |  |  |  |
| adjusted for CR + NANA^c^ | 1.58 (1.15–2.16) | 3.92 (2.47–6.29) | NA | NA |
| **Overall survival, all cases, HR high vs low CPG (95% CI)** |  |  |  |  |
| Adjusted^d^ | 1.53 (1.21–1.94) | 1.42 (0.96–2.11) | NA | NA |

All cases versus all controls (NCI-Maryland 375 vs 471; Colorado 286 vs 218 with complete covariates). Odds ratios are per standard deviation of the within-study standardized log_10_ CPG (nM); pooled models include a study term, and heterogeneity was assessed by Cochran’s *Q* test. CI, confidence interval; CR, creatine riboside; HR, hazard ratio; NA, not applicable; NANA, *N*-acetylneuraminic acid; OR, odds ratio; SD, standard deviation. ^a^Adjusted for age, sex, race and smoking status (never vs ever). ^b^P value for the CPG × smoking-status interaction term in the adjusted model. ^c^CPG and CR + NANA each dichotomized at the study-specific median, in a model adjusted for age, sex, race and smoking status. ^d^Cases with complete staging (NCI-Maryland 364 cases, 288 deaths; Colorado 284 cases, 103 deaths), adjusted for stage, age, sex and smoking status. All tests were two-sided.

**Table S3** Sensitivity analysis excluding four Colorado participants with uncertain case–control classification

| **Analysis** | **Main analysis** | **Sensitivity analysis** |
| --- | --- | --- |
| Colorado participants, cases / controls | 286 / 219 | 284 / 217 |
| Colorado never-smokers, cases / controls | 80 / 108 | 79 / 108 |
| Colorado, all participants, adjusted OR per SD^a^ | 1.91 (1.52–2.39) | 1.88 (1.50–2.36) |
| Colorado, never-smokers, adjusted OR per SD^b^ | 1.81 (1.24–2.65) | 1.80 (1.23–2.63) |
| Pooled, never-smokers with stage I–II disease, adjusted OR per SD^b^ | 1.38 (1.04–1.82) | 1.37 (1.04–1.81) |
| Colorado, never-smoker cases, unadjusted HR for high vs low CPG | 2.15 (1.00–4.64) | 2.29 (1.06–4.94) |

Estimates are shown with 95% confidence intervals. The four excluded participants (two cases, one of them a never-smoker, and two ever-smoker controls) had uncertain case–control classification in the study records. HR, hazard ratio; OR, odds ratio; SD, standard deviation. ^a^Adjusted for age, sex, race and smoking status. ^b^Adjusted for age, sex and race; the pooled model includes a study term.

**Supplementary Figures**

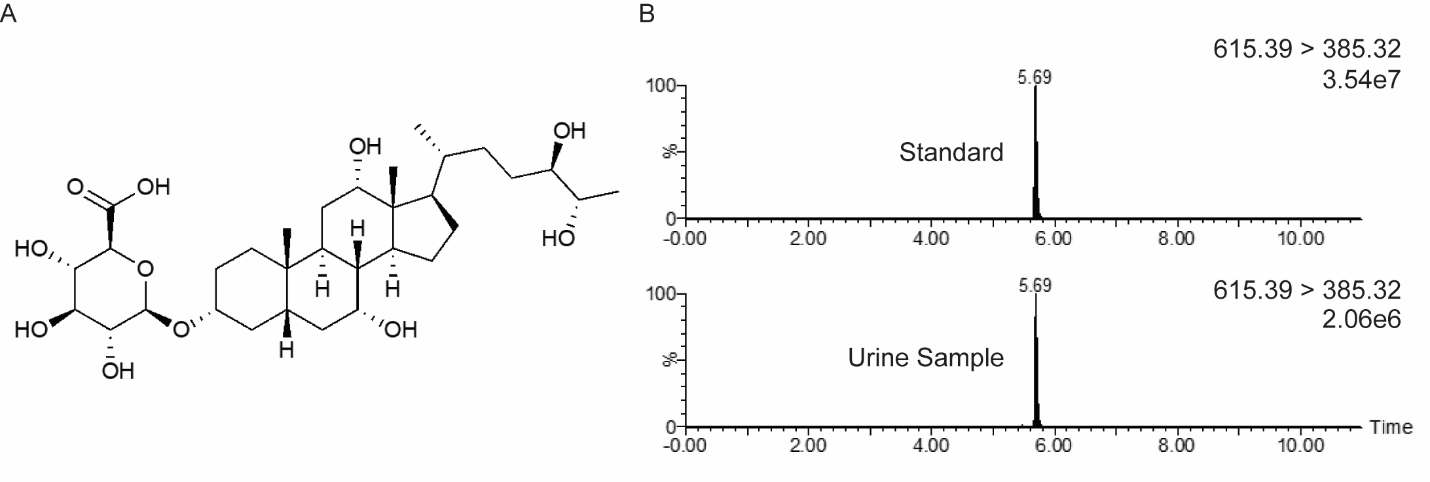

**Figure S1 Structural identification and chromatographic detection of urinary CPG.** **A** Chemical structure of 27-nor-5β-cholestane-3α,7α,12α,24*R*,25*S*-pentol glucuronide (CPG). **B** Multiple-reaction-monitoring chromatograms (615.39 → 385.32) for the synthesized authentic CPG standard (top) and a representative urine sample (bottom), confirming identity by matched retention time (5.69 min).

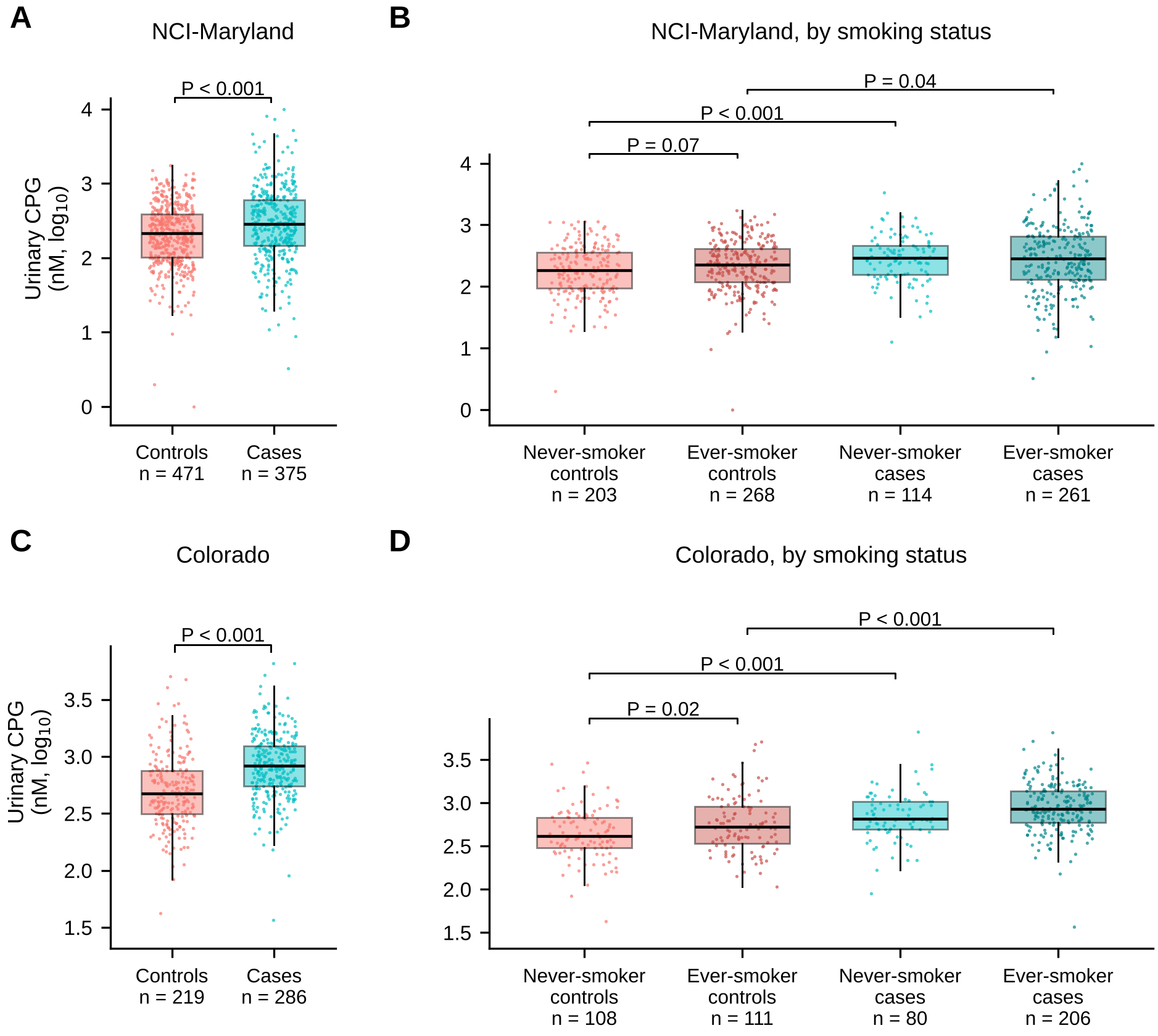

**Figure S2 Urinary CPG in all participants, overall and by smoking status.** Urinary CPG (nM, log_10_ scale) in controls and non-small cell lung cancer cases in the **A** NCI-Maryland and **C** Colorado studies, and in never-smoker and ever-smoker controls and cases in the **B** NCI-Maryland and **D** Colorado studies. Boxes show the median and interquartile range, whiskers extend to 1.5 times the interquartile range, and individual participant values are overlaid. Group sizes are given beneath each box. P values are from two-sided Mann–Whitney *U* tests; in **B** and **D** they are Holm-adjusted for four prespecified pairwise comparisons, of which three are shown (ever-smoker vs never-smoker cases: P = 0.98 in NCI-Maryland and P = 0.008 in Colorado). Vertical scales differ between studies because samples were analyzed in separate batches.

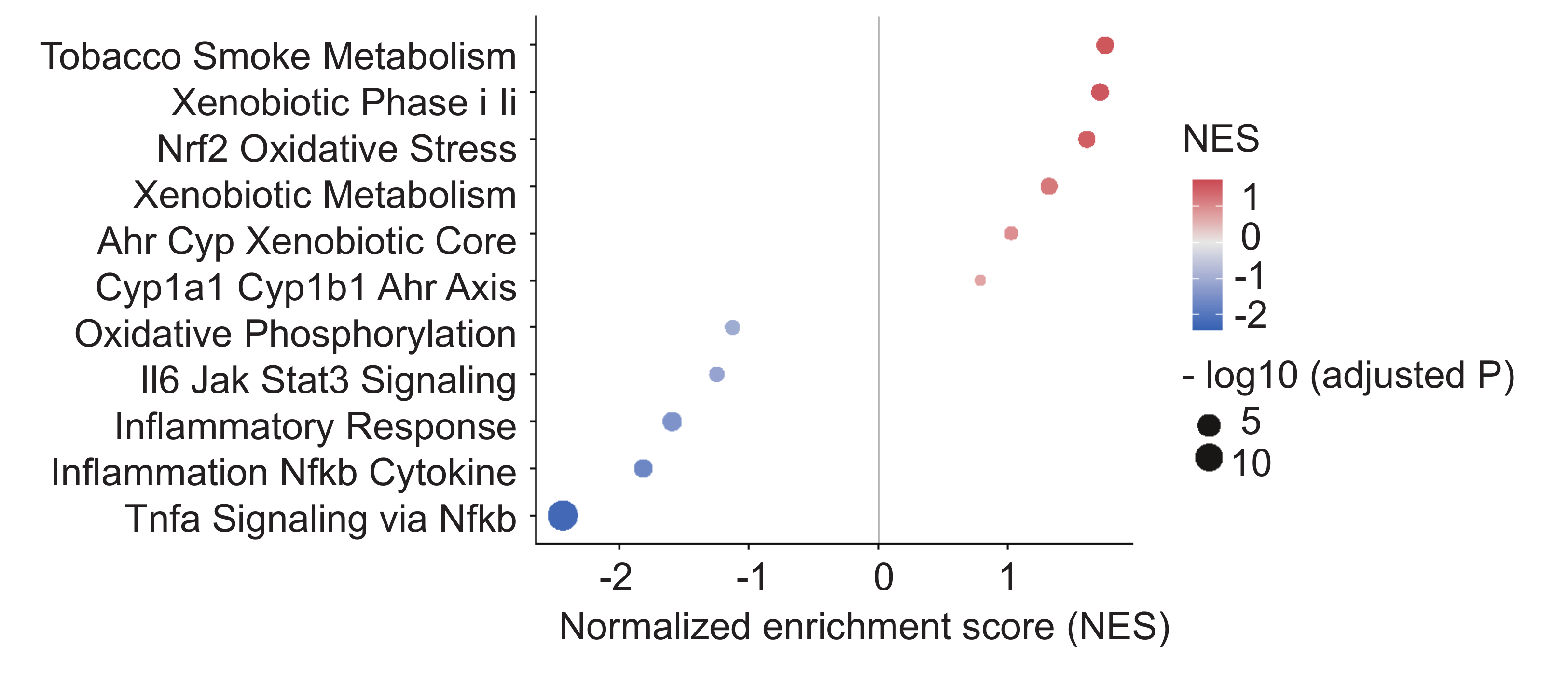

**Figure S3 Tumor gene-set enrichment by median-split urinary CPG.** Gene-set enrichment analysis of tumors from patients with high versus low urinary CPG (median split), against a curated collection of xenobiotic-metabolism, AhR/CYP, Nrf2 oxidative-stress and inflammatory gene sets. Dot color indicates the normalized enrichment score (NES); dot size indicates −log_10_ Benjamini–Hochberg-adjusted P. In the same median-split analysis, the Hallmark P53 pathway, apoptosis, hypoxia and epithelial–mesenchymal transition sets were negatively enriched (NES −1.6 to −3.2), but these associations were not reproduced with CPG modeled as a continuous variable (Figure S4A). Never-smokers contributed only a small minority of sequenced tumors.

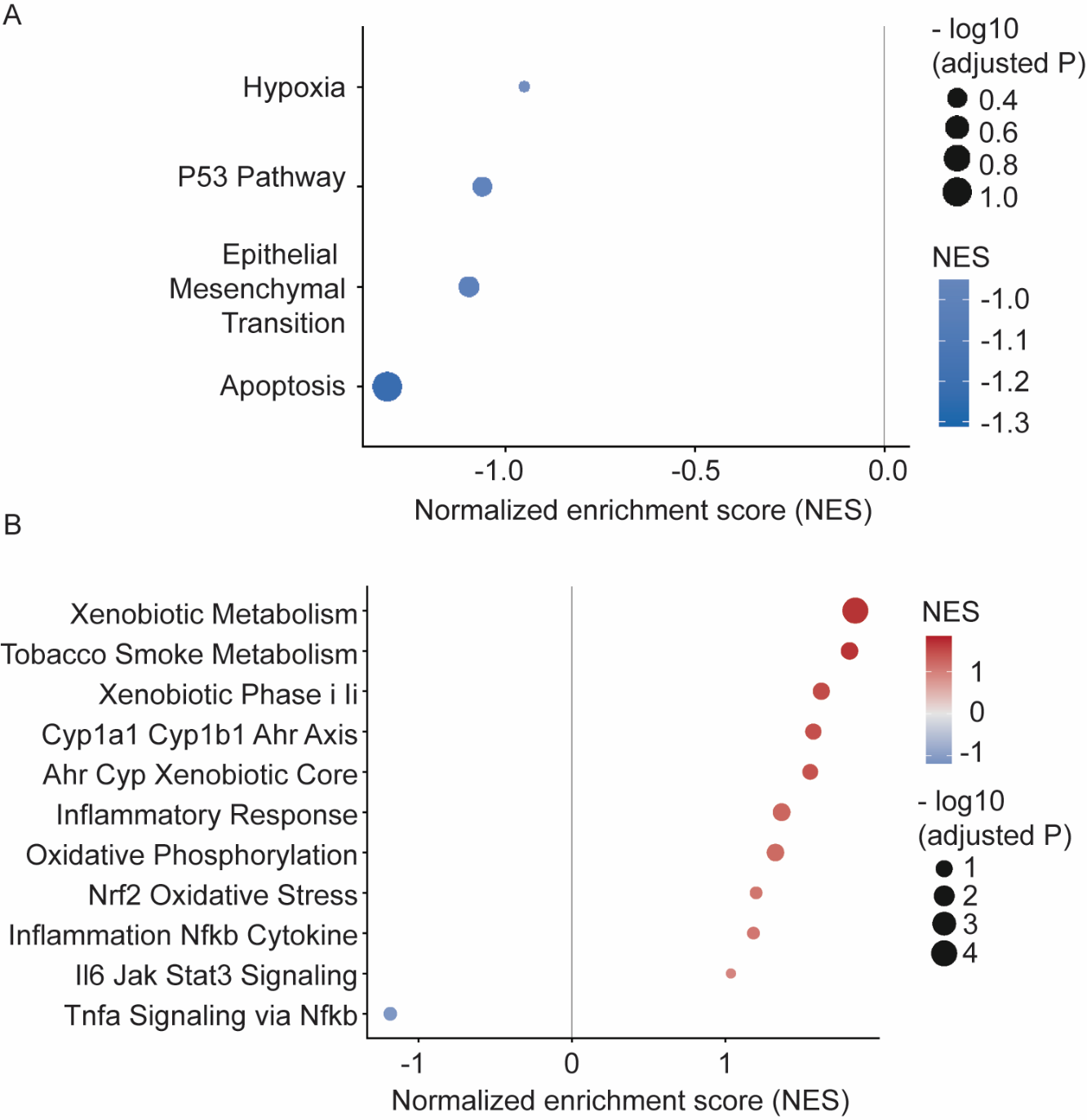

**Figure S4 Tumor gene-set enrichment with urinary CPG as a continuous variable.** **A** MSigDB Hallmark sets: hypoxia, P53 pathway, epithelial–mesenchymal transition and apoptosis are negatively enriched but with NES of only −0.9 to −1.3 and no adjusted P below 0.1. **B** Curated sets: the xenobiotic-metabolism, tobacco-smoke-metabolism, xenobiotic phase I/II, CYP1A1/CYP1B1 AhR axis, AhR/CYP core and Nrf2 oxidative-stress sets are positively enriched, reproducing the median-split analysis (Figure S3); the oxidative-phosphorylation and inflammatory sets change direction between analyses and are not interpreted.

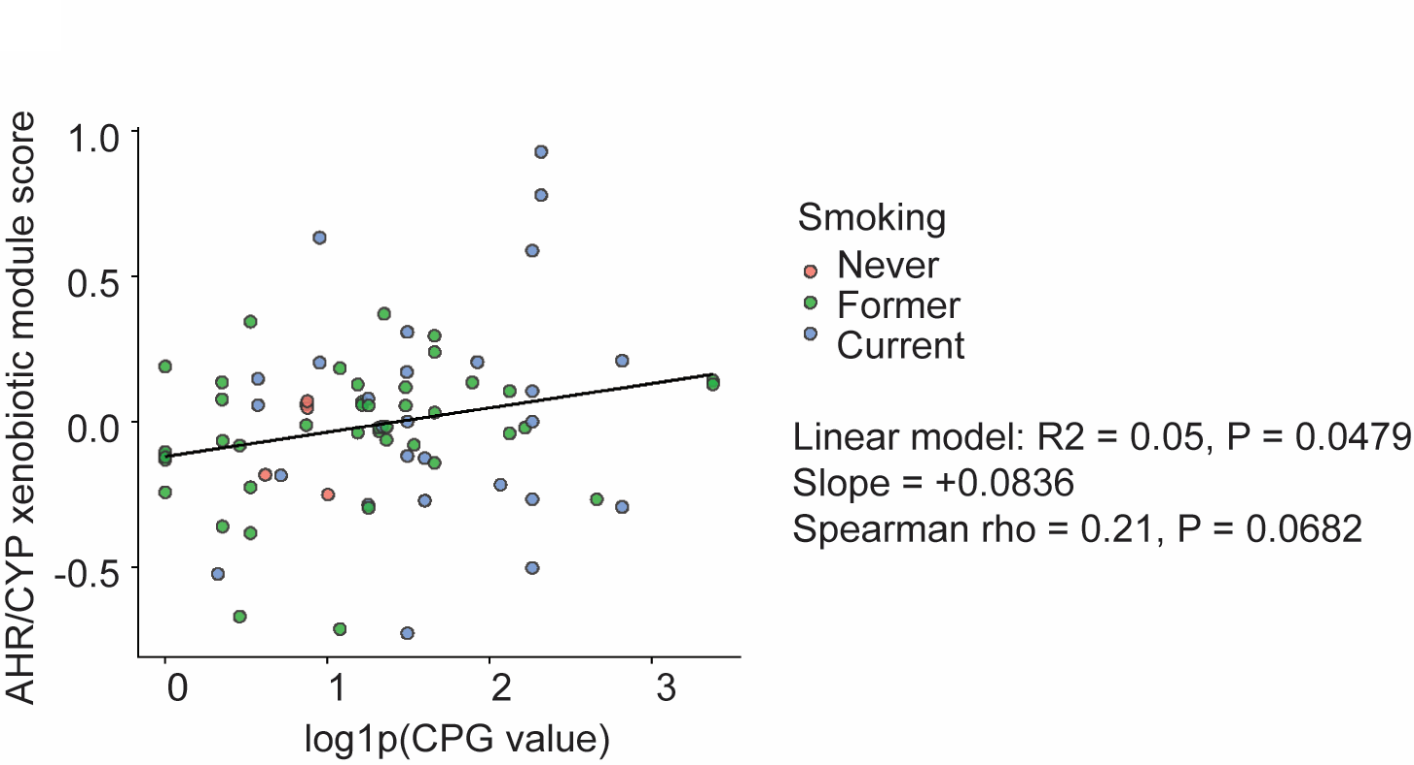

**Figure S5 AhR/CYP xenobiotic module score versus urinary CPG.** Module score (mean *z*-scored expression of member genes) plotted against log1p-transformed urinary CPG across the RNA-sequencing cohort, colored by smoking status. Linear model *R*^2^ = 0.05, P = 0.048; Spearman ρ = 0.21, P = 0.07. Never-smokers contribute only a small minority of samples.
